# Comparative quantitative analysis of SARS- CoV-2 Spike neutralizing antibody titers following two anti COVID-19 vaccines in India

**DOI:** 10.1101/2021.08.28.21262753

**Authors:** GV Chanukya, Aparna Srikantam

## Abstract

In COVID 19 Pandemic,first line of defence is effective vaccination program.Because of multiple platforms available for vaccine production we tested relative immunogenicity of two vaccines available in India, Covaxin® and Covishield® We performed quantitative analysis of neutralizing antibodies to SARS Cov2 spike (receptor binding domain) protein, from sera of 53 subjects who completed vaccines schedules.There was significantly higher immunogenic response with Covishield® as compared to Covaxin® and are independent of age. Studies on a large scale with long term follow up are needed to further advance the knowledge in this domain.

## Introduction

Ever since SARS-CoV-2 spread globally, since 2019,there have been herculean efforts by many countries to produce effective vaccines against the organism. WHO collaborated on these efforts in the name of COVAX for global equitable access to COVID 19 Vaccines^1^. In India as of August 2021, two vaccines have been extensively utilized. Covaxin® is an inactivated virus-based COVID-19 vaccine developed by Bharat Biotech in collaboration with the Indian Council of Medical Research^2^. Second one is The Oxford–AstraZeneca COVID-19 vaccine, sold under the brand name Covishield®, is a viral vector vaccine^3^. Both are administered by two doses of intramuscular injections and have good safety profiles. effective against SARS-CoV-2 ariants against delta variants. All these studies were done in different backgrounds of populations and there were no head to head trials comparing either their efficacy or effectiveness against emerging variants. While most of the studies were conducted as part of the vaccine trials, there is a paucity of information on how the responses were comparable between the two vaccines.In many studies titres of IgG is regarded as protective immunity against the virus^4,5^. Such information would facilitate improving immunoprophylactic strategies for SARSCoV2. With this background, we sought to compare the protective efficacy of Covaxin® and Covishield® vaccines through quantitative analysis of antibody titres response at least 10 days (Range 10 – 90 days) after completing the full schedule.

## Materials and methods

We collected data from 54 patients attending Dr Chanukya Endocrinology clinic,Hyderabad,India over a period of two months who had successfully completed schedule of two dose series of both vaccines. We chose a cut off of 10 days post second dose to avoid assay interference with natural infection in the interval. Demographics including age,gender,past incidence of natural covid infection and its interval and whether hospitalization was required or not were collected. We quantified serum IgG anti spike neutralizing antibodies directed for receptor binding domain (RBD), employing CLIA Any numeric value over and above 250 was rounded off to 250. We hypothesized the higher value, greater immunogenic response.Hence we segregated subjects into two groups viz. group 1 IgG titers ≥ 250 and < 250. We excluded one patient for missing IgG numeric value.

## Results

Of the 53 subjects (males 30 and females 23), 28 have titers ≥ 250. Their mean age in years was 55.32 ± 7.5. Seven had a past history of COVID 19 disease. The mean age in years of the second group (IgG titers < 250, n= 25) was 56.96 ± 14.56. Among them three had a history of COVID 19 disease. For assessing relative efficacy of two vaccines we employed two proportion test, null hypothesis being both are equally efficacious. P value is statistically significant when it is < 0.05.Fisher exact value was 0.029. So the proportion of persons having Ig G titers > 250 for the two vaccines are different (Covishield® produced a more immunogenic response compared to Covaxin®). Also we employed two sample t-test for estimating means of IgG titers of respective vaccines. For Covaxin® it was 23.7 ± 50.05 and for Covishield® it was 126 ± 149 .p value was 0.04 (p < 0.05 being statistically significant). suggesting there is a significant difference in mean IgG values.

## Discussion

Present study highlights preliminary findings on the variable efficacy of the two widely used SARS-CoV-2 vaccines in India ^6^.As an inactivated vaccine, Covaxin® uses a more traditional technology that is similar to the inactivated polio vaccine. SARS-CoV-2 isolated by India’s National Institute of Virology was used to grow using a vero cell line. Then virions are deactivated with beta-propiolactone. The resulting inactivated whole virions are then mixed with the aluminium-based adjuvant Alhydroxiquim-II. In a double blind randomised phase III control trial,, overall vaccine efficacy of 77·8% (95% CI: 65·2–86·4) was noted It conferred 65·2% (95% CI: 33·1–83·0) protection against the SARS-CoV-2 Variant of Concern, B.1.617.2 (Delta)^7^. On other hand Covishield® was developed by Oxford University and AstraZeneca, using the modified chimpanzee adenovirus ChAdOx1 as a vector. Efficacy of the vaccine is % at preventing symptomatic COVID-19 beginning at 22 days following the first dose and 81.3% after the second dose^8^. For symptomatic COVID-19 infection after the second dose, the vaccine is 81% effective against the Alpha variant (lineage B.1.1.7), and 61% against the Delta variant (lineage B.1.617.2)^9^.

As it is already known the vaccine trials of Covishield® have been completed before its use for community vaccination, while the Covaxin® has been put into use much before the full results of the clinical trials are published in peer reviewed journals. Both of course were mentioned for emergency use meaning that the manufacturers would continue to improve the product for its best purpose. However, given that both the vaccines are being used widely across India, it is logical to know how these two vaccines are performing in the real world, as compared to their efficacy during the trials. The trials were mostly based on measuring the protection against natural infection since it will not be possible to wait for months to measure the antibody titre. In this scenario it is logical to generate data on immunogenicity of each of these vaccines in order to understand their behaviour and predict the protective efficacy vis a vis the prevailing SARS-CoV-2 variants. This study attempted bringing out such information on a pilot basis. We found that IgG titres were significantly greater with Covisheld ®compared to Covaxin®. Effects of age, gender,duration of post vaccination for the initial three months are not significant.

## Conclusion

Our pilot findings suggest greater immunogenicity of Covishield® compared to Covaxin ®while warranting further studies with large representative samples across the country which may further include other vaccines such as Sputnik® as well.

## Data Availability

All the data that is referred in the manuscript is available.

## References

1. COVAX-Working for equitable access to COVID vaccines. Available online at https://www.who.int/initiatives/act-accelerator/covax (Accessed on August 26-08-2021)

2. COVAXIN® - India’s First Indigenous COVID-19 Vaccine. Available online at https://www.bharatbiotech.com/covaxin.html (Accessed on August 26-08-2021)

3. ChAdOx1 nCoV- 19 Corona Virus Vaccine (Recombinant) COVISHIELDTM. Available online at https://www.seruminstitute.com/product_covishield.php (Accessed on August 26-08-2021)

4. David S. Khoury, Deborah Cromer et al. Neutralizing antibody levels are highly predictiveof immune protection from symptomatic SARS-CoV-2 infection. Nat Med 27, 1205–1211 (2021). https://doi.org/10.1038/s41591-021-01377-8

5. Yamayoshi S, Yasuhara A et al. Antibody titers against SARS-CoV-2 decline, but do notdisappear for several months. EClinicalMedicine. 2021 Feb;32:100734. doi: 10.1016/j.eclinm.2021.100734. Epub 2021 Feb 11. PMID: 33589882; PMCID: PMC7877219.

6. Awadhesh K, Singh AK et al. Antibody Response after Second-dose of ChAdOx1-nCOV (Covishield™®) and BBV-152 (Covaxin™®) among Health Care Workers in India: Final Results of Cross-sectional Coronavirus Vaccine-induced Antibody Titre (COVAT) study. MedRxiv 2021.06.02.21258242; doi: https://doi.org/10.1101/2021.06.02.21258242

7. Raches Ella, Krishna Mohan Vadrevu et al. Safety, and lot to lot immunogenicity of aninactivated SARS-CoV-2 vaccine (BBV152): a, double-blind, randomised, controlled phase 3 trial. medRxiv 2021.06.30.21259439; doi:https://doi.org/10.1101/2021.06.30.21259439

8. Merryn Voysey, Sue Ann Costa Clemens et al. Oxford COVID Vaccine Trial Group. “Single-dose administration and the influence of the timing of the booster dose on immunogenicity and efficacy of ChAdOx1 nCoV-19 (AZD1222) vaccine: a pooled analysis of four randomised trials”. Lancet. 397 (10277): 881–891. doi:10.1016/S0140-6736(21)00432-3. PMC 7894131. PMID 33617777.

9. Sheikh A, McMenamin J et al. “SARS-CoV-2 Delta VOC in Scotland: demographics, riskof hospital admission, and vaccine effectiveness”. The Lancet. 397 (10293). Table S4. doi:10.1016/S0140-6736(21)01358-1. ISSN 0140-6736. PMC 8201647. PMID 34139198.

